# Clinical characteristics of 51 patients discharged from hospital with COVID-19 in Chongqing,China

**DOI:** 10.1101/2020.02.20.20025536

**Authors:** Lei Liu, Jian-Ya Gao, Wan-mei Hu, Xian-xiang Zhang, Lian Guo, Chun-qiu Liu, Yue-wu Tang, Chun-hui Lang, Fang-zheng Mou, Zheng-jun Yi, Qin-qin Pei, Kai Sun, Jiang-lin Xiang, Jiang-feng Xiao

**Author notes:** Corresponding author: Lei Liu. Department of Nephrology,Chongqing Three Gorges Central Hospital,Chongqing University Three Gorges Hospital,165Xin Cheng Road, Wan zhou District, Chongqing, China. Jian-Ya Gao and Lei Liu contributed equally to this article.

## Abstract

**Background:** Since December 2019, Severe acute respiratory syndrome coronavirus 2(SARS-CoV-2)-infected disease (Coronavirus Disease 2019,COVID-19) emerged in Wuhan, China,and rapidly spread throughout China,even throughout the world. We try to describe the epidemiological and clinical characteristics of COVID-19 in non-Wuhan area,and explore its effective treatment.

**Methods:** Retrospective, single-center case series of the 51 hospitalized patients with confirmed COVID-19 at Chongqing University Three Gorges Hospital in Chongqing, China, from January 20 to February 3, 2020;The discharge time was from January 29 to February 11, 2020. The main results and indicators of epidemiology, demography, clinical manifestation, laboratory examination, imaging data and treatment data of 51 patients with covid-19 were collected and analyzed. The changes of blood routine and biochemical indexes at discharge and admission were compared. Compare the clinical characteristics of severe patients (including severe and critical patients) and non-severe patients (general patients).

**Results:** Of 51 hospitalized patients with COVID-19, the median age was 45 years (interquartile range, 34-51; range, 16-68 years) and 32 (62.7%) were men.43(84.3%) patients had been to Wuhan or Other Hubei areas outside Wuhan,and 4(7.7%) patients had a clear contact history of COVID-19 patients before the onset of the disease, and 4 (7.7%) patients had no clear epidemiological history of COVID-19.Common symptoms included fever (43 [84.3%]), cough (38 [74.5%]) and fatigue (22 [43.1%]). Lymphopenia was observed in 26 patients (51.0%), and elevated C-reactive protein level in 32 patients (62.7%). Ground-glass opacity was the typical radiological finding on chest computed tomography (41 [80.4%]),Local consolidation of pneumonia in some patients(17 [33.3%]).Most of the patients were treated with traditional Chinese medicine decoction (28 [54.9%]),all of them received aerosol inhalation of recombinant human interferon a-1b for injection and oral antiviral therapy with Lopinavir and Ritonavir tablets (51 [100%]); Most of the patients were given Bacillus licheniformis capsules regulated intestinal flora treatment (44 [86.3%]). 10 patients (19.6%) received short-term (3-5 days) glucocorticoid treatment. Compared with non-severe patients (n = 44), severe patients (n = 7) were older (median age, 52 years vs 44 years), had a higher proportion of diabetes mellitus (4 [57.1%] vs 0 [0.0%]), most of them needed antibiotic treatment (7 [100%] vs 4 [9.1%], most of them needed nutritional diet (6 [85.7%) vs 0 [0.0%], and were more likely to have dyspnea (6 [85.7%] vs 5 [11.4%]),most of them needed noninvasive mechanical ventilation (6 [85.7%] vs 0 [0.0%]). Except one patient died, the remaining 50 patients were discharged according to the discharge standard, the common clinical symptoms disappeared basically, the lymphocyte increased significantly (P=0.008), CRP decreased significantly (P <0.001). The median length of stay was 12 days (IQR, 9-13).

**Conclusion:** In 51 single center cases confirmed as COVID-19 and discharged from the hospital, 13.7% of the patients were severe. The main clinical symptoms of patients with COVID-19 were fever, cough and asthenia,Some patients had obvious dyspnea. They had clinical laboratory and radiologic characteristics. There is no specific drug treatment for the disease. For the treatment of COVID-19, in addition to oxygen inhalation and antiviral treatment, attention should be paid to the dialectical treatment of traditional Chinese medicine, regulation of intestinal flora, nutritional support treatment and other comprehensive treatment.

## Introduction

In December 2019, a group of acute respiratory diseases with unknown causes[1], now known as Coronavirus Disease 2019(COVID-19), caused by Severe acute respiratory syndrome coronavirus 2(SARS-CoV-2)[2-4],occurred in Wuhan, Hubei Province, China. The 2019 novel coronavirus (2019-nCoV) was declared a Public Health Emergency of International Concern(PHEIC)by the world health organization on January 30, 2020[5],.The disease has spread rapidly from Wuhan to all over China and other countries [5-9].In China(excluding Hong Kong, Macao and Taiwan),As of 24:00 on February 14, 2020, 66492 confirmed cases have been reported, 8096 discharged cases have been cured and 1523 deaths have been reported [10].

Despite the rapid spread worldwide,the clinical characteristics of COVID-19 remain largely unclear. Recently, a series of studies have reported the epidemiological and clinical characteristics of patients with COVID-19[1,11-14].In three recent studies documenting the clinical manifestations of 41, 99and138 patients respectively with laboratory-confirmed COVID-19 who were admitted to Wuhan, the severity of some cases with COVID-19 mimicked that of severe acute respiratory syndrome coronavirus (SARS-Cov) [[1,11-12].Recently, a large sample study recorded data of 1099 laboratory confirmed cases, According to the researchers [13]; The SARS-CoV-2 epidemic spreads rapidly by human-to-human transmission. The disease severity(including oxygen saturation, respiratory rate, blood leukocyte/lymphocyte count and chest X-ray/CT manifestations) predict poor clinical outcomes. Another large sample study suggests[14],Compared with SARS-CoV, 2019-nCoV had comparable transmissibility and lower case fatality rates (CFR). Their findings based on individual-level surveillance data emphasize the importance of early detection of elderly patients, particularly males, before symptoms progress to severe pneumonia.

At present, many studies are aimed at the patients with COVID-19 in or including Wuhan area, Hubei Province. We know that [13-14], the medical resources in Wuhan area are in short supply in the early stage, and it may take a long time from the onset of the patients to the diagnosis, and the patients admitted to the hospital in the early stage tend to be in a serious condition. As of 24:00 on February 14, 2020, the mortality rate in Wuhan area is 2.96% (The mortality rate was calculated by the cumulative number of deaths / the number of confirmed cases involved), Significantly higher than 1.40% in non Wuhan areas of China (excluding Hong Kong, Macao and Taiwan) and 0.55% in non Hubei areas of China (excluding Hong Kong, Macao and Taiwan)[10].

At present, there is no effective treatment or vaccine for COVID-19.Therefore, it is necessary to explore the epidemiological and clinical characteristics of COVID-19 patients hospitalized in non-hubei areas.

The Chongqing University Three Gorges Hospital is one of the four designated hospitals in Chongqing for the treatment of COVID-19 patients, mainly for the treatment of COVID-19 patients in the northeast of Chongqing. As of 24:00 on February 14, a total of 537 confirmed cases and 5 deaths have been reported in Chongqing [15].As of February 11, 2020, Chongqing University Three Gorges Hospital has involved 50 cases of cured discharge and 1 case of death. Here, by collecting the data of 51 laboratory confirmed cases, we try to provide a description of the epidemiology and clinical characteristics of COVID-19 patients in Chongqing, and explore the changes of clinical characteristics of patients during hospitalization.

## Data and Methods

### Case data

51 patients were admitted to the Chongqing University Three Gorges Hospital from January 20, 2020 to February 3, 2020. All of them were confirmed cases. The discharge time was from January 29, 2020 to February 11, 2020

## Methods

### Data sources

This case series was approved by the institutional ethics board of Chongqing University Three Gorges Hospital. All patients diagnosed as COVID-19 in the Chongqing University Three Gorges Hospital, who had been discharged by February 11, 2020, were included in the study. Oral consent was obtained from patients,Written informed consent was waived in light of the urgent need to collect clinical data. All COVID-19 patients were diagnosed according to World Health Organization interim guidance[16] and ⟪Diagnosis and treatment of novel coronavirus pneumonia (trial version fifth) ⟫ [17]. Laboratory confirmation of the SARS-CoV-2 was achieved through the Chongqing Wanzhou Center for Disease Control and Prevention.

### Data Collection

The internal medicine research group of the Chongqing University Three Gorges Hospital analyzed the patient’s medical records. Epidemiological, clinical, laboratory and radiologic characteristics, as well as treatment and outcome data are obtained through electronic medical records. The data were collected and reviewed by a team of trained doctors. The recorded information includes demographic data, medical history, exposure history, potential comorbidities, symptoms, physical signs, laboratory test results (including blood cell analysis, liver and kidney function, coagulation test, C-reactive protein, procalcitonin, lactate dehydrogenase, creatine kinase,Interleukin 6 and urine routine, etc.), chest computed tomography (CT) and treatment measures (i.e. antiviral treatment, corticosteroid treatment, respiratory support, traditional Chinese medicine treatment, nutritional support, etc.). The onset date is defined as the date when the symptom first appears. The severity of COVID-19 was classified according to World Health Organization interim guidance[16] and ⟪Diagnosis and treatment of novel coronavirus pneumonia (trial version fifth) ⟫ [17]. Acute respiratory distress syndrome(ARDS)was defined according to the Berlin definition[18]. The durations from onset of disease to diagnosis,diagnosis to discharge were recorded.

### Statistical analysis

Continuous variables were expressed as the medians and interquartile ranges (IQR) as appropriate. Categorical variables were summarized as the counts and percentages in each category. Means for continuous variables were compared using independent group t tests when the data were normally distributed; otherwise, the Mann-Whitney test was used. Proportions for categorical variables were compared using the χ 2test, although the Fisher exact test was used when the data were limited. All statistical analyses were performed using SPSS (Statistical Package for the Social Sciences) version 19.0 software (SPSS Inc). For unadjusted comparisons, a 2-sided α of less than 0.05 was considered statistically significant. The analyses have not been adjusted for multiple comparisons and, given the potential for type I error, the findings should be interpreted as exploratory and descriptive.

## Results

### General baseline data

The median age of 51 inpatients with COVID-19 was 45 years (interquartile range, 34-51; range, 16-68 years), including 32 males (62.7%) and 19 females (37.3%); There were 8 cases (15.7%) with comorbidities, 4 cases (7.8%) with diabetes, 4 cases (7.8%) with hypertension, 1 case (2.0%) with diabetes and hypertension, 1 case (2.0%) with chronic hepatitis B and 1 case (2.0%) with schizophrenia (Table 1). Of the 51 patients, 44 (86.3%) were non severe (common type), 7 (13.7%) were severe (4 [7.8%] were severe, 3 [5.9%] were critical).

**Table 1.** Clinical characteristics of 51 patients with COVID-19.

|  | N0.(%) |  |  | P value |
| --- | --- | --- | --- | --- |
|  | Total<br>(N=51) | Non-severe<br>(N=44) | Severe<br>(N=7) |  |
| Age, median(IQR),Y | 45 ( 34-51 ) | 44 ( 33-49 ) | 52 ( 44-60 ) | 0.009 |
| Sex |  |  |  |  |
| female | 19 ( 37.3 ) | 16 ( 36.4 ) | 3 ( 42.9 ) | 0.109 |
| male | 32 ( 62.7 ) | 28 ( 63.7 ) | 4 ( 57.1 ) |  |
| Comorbidities |  |  |  |  |
| Hypertension | 4 ( 7.8 ) | 3 ( 6.8 ) | 1 ( 14.3 ) | 0.457 |
| Diabetes | 4 ( 7.8 ) | 0 ( 0 ) | 4 ( 57.1 ) | <0.001 |
| Chronic hepatitis B* | 1 | 0 ( 0 ) | 1 ( 14.3 ) | 0.137 |
| Schizophrenia<sup>△</sup> | 1 | 1 ( 2.3 ) | 0 ( 0 ) | 1.0 |
| Signs and symptoms |  |  |  |  |
| dyspnea | 11 ( 21.6 ) | 5 ( 11.4 ) | 6 ( 85.7 ) | <0.001 |
| fever | 43 ( 84.3 ) | 37 ( 84.1 ) | 6 ( 85.7 ) | 0.913 |
| cough | 38 ( 74.5 ) | 32 ( 72.7 ) | 6 ( 85.7 ) | 0.464 |
| expectoration | 16 ( 31.4 ) | 14 ( 31.8 ) | 2 ( 28.6 ) | 0.863 |
| pharyngalgia | 2 ( 3.9 ) | 2 ( 4.5 ) | 0 ( 0 ) | - |
| muscular soreness | 6 ( 11.8 ) | 5 ( 11.4 ) | 1 ( 14.3 ) | 1.0 |
| A stuffy or runny nose | 3 ( 5.9 ) | 1 ( 2.3 ) | 2 ( 28.6 ) | 0.046 |
| diarrhea | 4 ( 7.8 ) | 4 ( 9.1 ) | 0 ( 0 ) | 1.0 |
| headache | 5 ( 9.8 ) | 5 ( 11.4 ) | 0 ( 0 ) | 1.0 |
| Dizziness | 7 ( 13.7 ) | 5 ( 11.4 ) | 2 ( 28.6 ) | 0.242 |
| nausea | 3 ( 5.9 ) | 3 ( 6.8 ) | 0 ( 0 ) | 1.0 |
| Retching or vomiting | 3 ( 5.9 ) | 3 ( 6.8 ) | 0 ( 0 ) | 1.0 |
| fatigue | 22 ( 43.1 ) | 16 ( 36.4 ) | 6 ( 85.7 ) | 0.014 |
| Diagnostic time, median(IQR),D | 6(3-8) | 5(3-8) | 6(6-9) | 0.359 |
| length of stay, median(IQR),D | 12(9-13) | 12(9-13) | 15(9-16) | 0.327 |

Comment : All data are presented as medians (interquartile ranges, IQR) and N0.(%).\*
Chronic hepatitis B denoted hepatitis B surface antigen tested positive, with or without elevated alanine or aspartate aminotransferase levels.^The case was stable disease. P values denoted the comparison between non-severe cases and severe cases. P<0.05 was considered statistically significant.

### Epidemiological data

43(84.3%)patients had been to Wuhan(40[78.4%])or Other Hubei areas outside Wuhan(3[5.9%]);(7.7%) patients had a clear contact history of COVID-19 patients before the onset of the disease, 3 patients had a clear contact history and contact date of covid-19 patients; 4 (7.7%)patients had no clear epidemiological history of COVID-19.The incubation period of 3 patients with definite contact history and contact date of COVID-19 patients was 2 days, 5 days and 5 days respectively. The median time from onset to diagnosis was 6 days (interquartile range, 3-8; range, 1-16 days) in 51 patients.

### Clinical characteristics

From onset to diagnosis,the common clinical symptoms were fever in 43 cases (84.3%);cough in 38 cases (74.5%), Most of them were dry cough with a small amount of expectoration in 16 cases (31.4%); asthenia in 22 cases (43.1%); dyspnea in 11 cases;the other uncommon clinical symptoms were: muscle pain in 6 cases (11.8%); pharyngeal pain in 2 cases (3.9%); nasal obstruction and / or runny nose in 3 cases (5.9%); dizziness in 7 cases (13.7%); headache in 5 cases (9.8%); diarrhea in 4 cases (7.8%); nausea in 3 cases (5.9%); retching or vomiting in 3 cases (5.9%)(Table 1).

2 patients (3.9%) developed diarrhea 1-2 days before fever, and 3 patients (5.9%) coughed 1-2 days before fever. Physical examination on admission,16 cases (31.4%) were found to have enlarged breath sounds in both lungs, and 4 cases (7.8%) could hear moist rales in both lungs. Diarrhea occurred in 13 patients (25.5%) during hospitalization. Asthenia, cough, muscle pain and other symptoms disappeared with the recovery of body temperature.

### Laboratory test results

The first day of admission,7 cases (13.7%) had WBC > 9.5 × 109 / L,6 cases (11.8%) had WBC < 3.5 × 109 / L, 11 cases (21.6%) had neutrophil count > 6.3 × 109 / L, 4 cases (7.8%) had neutrophil count < 1.8 × 109 / L, 26 cases (51.0%) had lymphocyte count < 1.1 × 109 / L, 10 cases (19.6%) had platelet count < 125 × 109 / L, 5 cases (9.8%) had AST > 40 U / L, 20 cases (39.2%) had LDH > 250 IU / L, 32 cases (62.7%) had CRP > 5 mg / L, 10 cases (19.6%) had D-dimer > 0.55 mg / L, 25 cases (49.0%) had procalcitonin > 0.046 ng / ml, 16 cases (31.4%) had IL-6 > 5.4pg/ml;8 cases (15.7%) were positive for routine urine protein, and all of them urine red blood cells were negative.

On the first day of admission (n = 51), the laboratory test results (leukocyte count, neutrophil count, lymphocyte count, hemoglobin, platelet, creatine kinase, lactate dehydrogenase, glutamic pyruvate transaminase, glutamic oxaloacetylase, albumin, creatinine, urea nitrogen, C-reactive protein, procalcitonin, interleukin-6) are shown in Table 2. There are many differences in laboratory test results between severe patients and non-severe patients (Table 2), including the significant decrease of lymphocyte count, the decrease of serum albumin level, and the significant increase of LDH, CRP, D-dimer and PCT levels.

**Table 2.** Laboratory findings of 51 patients with COVID-19.

|  | normal<br>value | Total<br>(N=51) | Non-severe<br>(N=44) | Severe<br>(N=7) | P<br>value |
| --- | --- | --- | --- | --- | --- |
| WBC | 3.5-9.5 | 5.4 ( 4.1-7.6 ) | 5.2 ( 4.1-7.6 ) | 5.7 ( 3.5-10.7 ) | 0.593 |
| N | 1.8-6.3 | 3.7 ( 2.5-6.0 ) | 3.5 ( 2.4-5.6 ) | 5.2 ( 3.2-9.9 ) | 0.865 |
| L | 1.1-3.2 | 1.1 ( 0.7-1.6 ) | 1.3 ( 0.9-1.7 ) | 0.37 ( 0.3-0.6 ) | <0.001 |
| HB | 120-160 | 136 ( 123-152 ) | 136 ( 123-152 ) | 132 ( 121-152 ) | 0.871 |
| PLT | 125-350 | 189 ( 134-235 ) | 192 ( 139-237 ) | 150 ( 116-225 ) | 0.423 |
| CK | 50-310 | 52 ( 38-79 ) | 59 ( 41-87 ) | 42 ( 35-46 ) | 0.333 |
| LDH | 120-250 | 231 ( 188-276 ) | 219 ( 182-264 ) | 299 ( 273-314 ) | 0.001 |
| AST | 15-40 | 21 ( 16-30 ) | 21 ( 16-29 ) | 29 ( 14-34 ) | 0.691 |
| ALT | 9-50 | 18 ( 14-30 ) | 18 ( 14-32 ) | 26 ( 10-30 ) | 0.694 |
| ALB | 40-55 | 40 ( 36-43 ) | 41 ( 38-44 ) | 35 ( 33-36 ) | 0.001 |
| Scr | 57-97 | 67 ( 60-79 ) | 68 ( 60-79 ) | 67 ( 50-68 ) | 0.283 |
| BUN | 3.1-8 | 4.1 ( 3.3-5.7 ) | 4.1 ( 3.1-5.6 ) | 5.1 ( 3.5-6.3 ) | 0.163 |
| D-D | 0-0.55 | 0.28 ( 0.19-0.51 ) | 0.28 ( 0.18-0.46 ) | 0.60 ( 0.28-1.40 ) | 0.014 |
| CRP | 0-5 | 10.5 ( 2.7-51.2 ) | 7.5 ( 2.4-23.8 ) | 135.7 ( 83.9-180.9 ) | <0.001 |
| PCT | <0.046 | 0.04 ( 0.03-0.07 ) | 0.04 ( 0.03-0.06 ) | 0.10 ( 0.05-0.24 ) | 0.004 |
| IL-6 | 0-5.4 | 0 ( 0-8.2 ) | 0 ( 0-7.3 ) | 4.6 ( 0-28.2 ) | 0.116 |

Comment : All data are presented as medians (interquartile ranges, IQR).Laboratory indicator
unit : HB g/l, CK U/L,LDH IU/L,AST U/L,ALT U/L,ALB g/l, Scr umol/l, BUN mmol/l, WBC 109/L,N 109/L,L 109/L,PLT 109/L,CRP mg/l,PCT ng/ml,IL-6 pg/ml, D-D mg/l.P values denoted the comparison between non-severe cases and severe cases. P<0.05 was considered statistically significant.

The results of laboratory test results of patients on the first day of admission (n = 50) and before discharge (n = 50) are compared (Table 3). We found that lymphocyte (L), PLT, CK, LDH and CRP of patients before discharge were significantly increased, while CK, LDH and CRP were significantly reduced (Table 3).

**Table 3.**
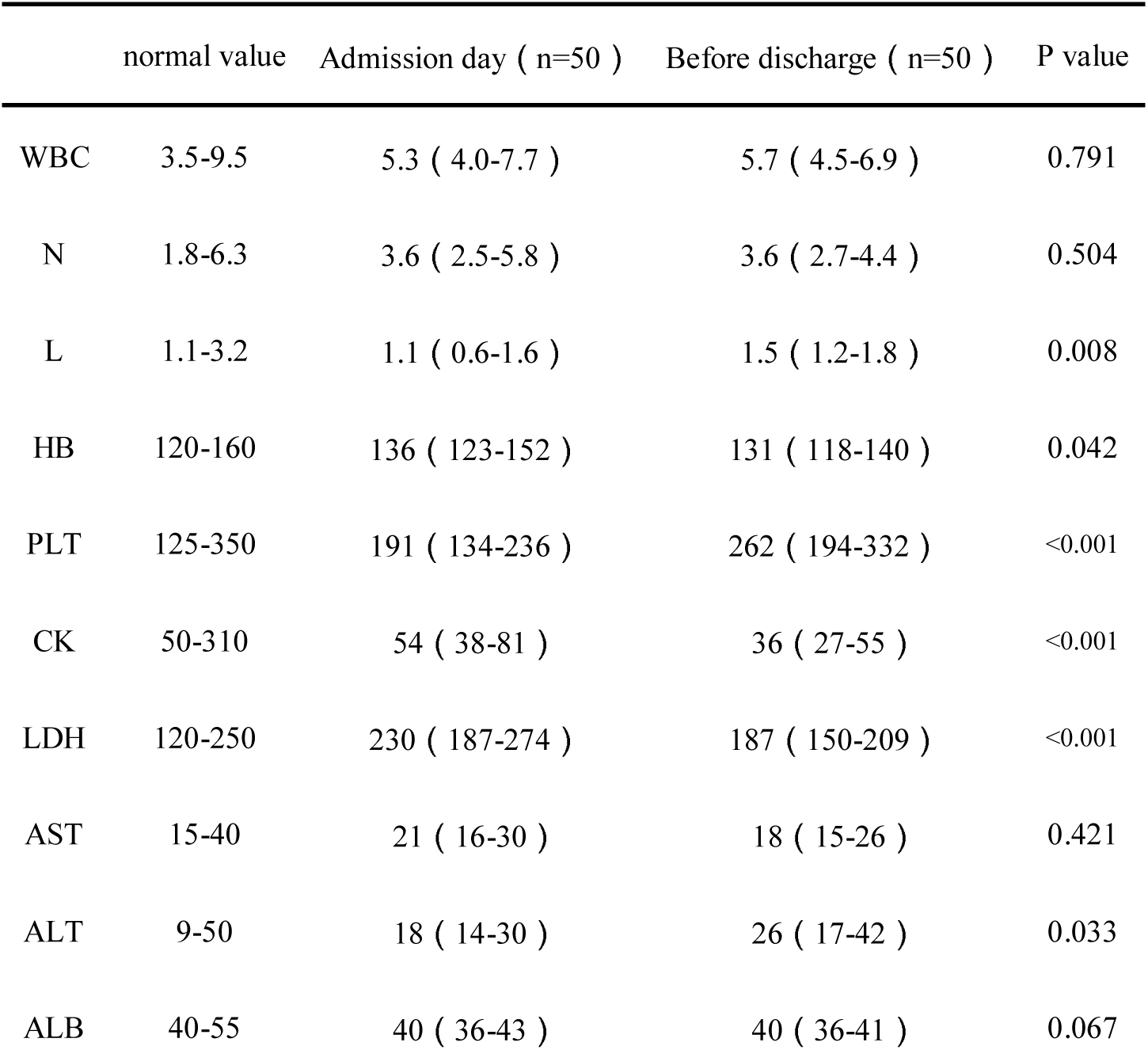
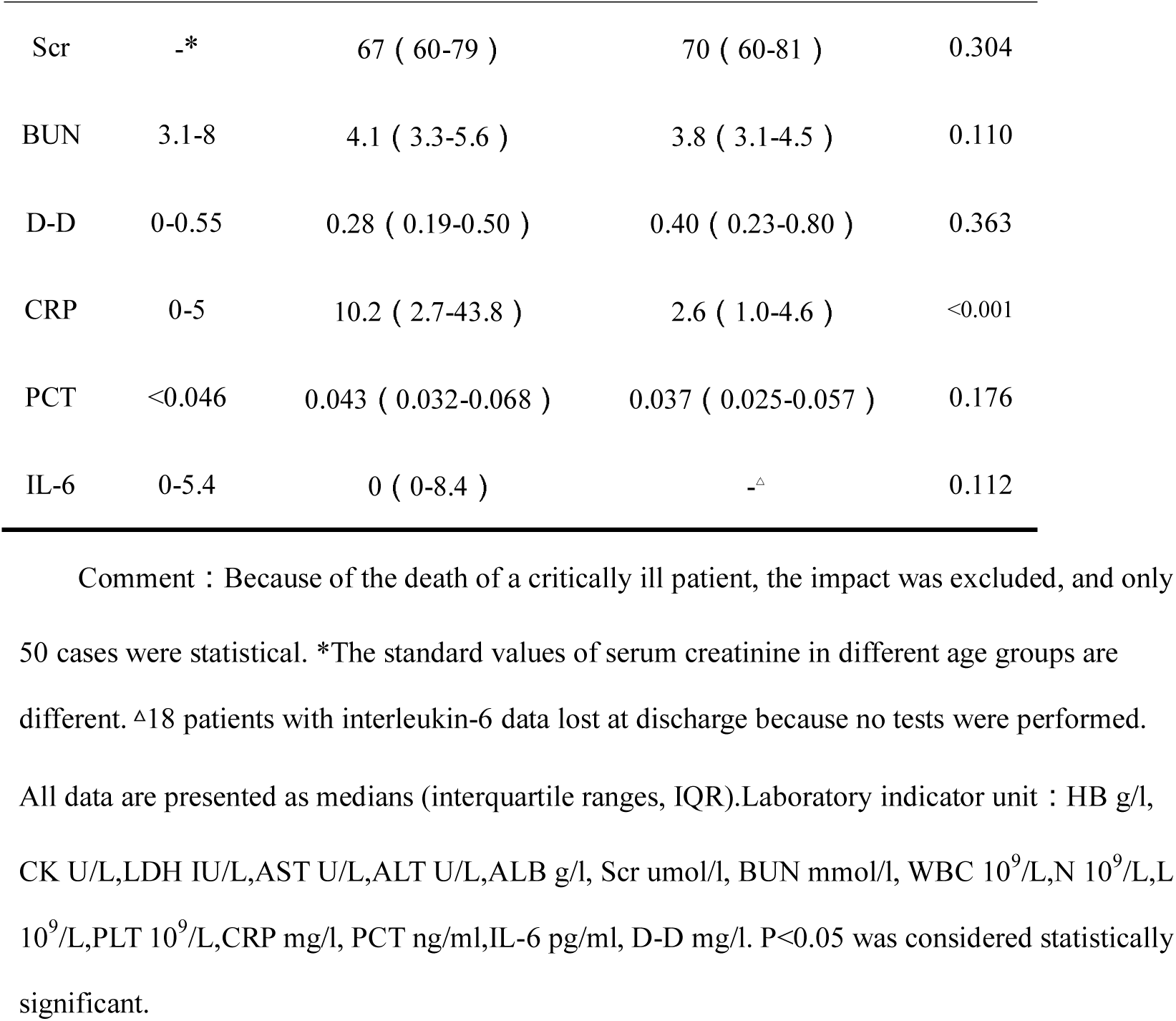
The comparison of laboratory findings of 50 patients with COVID-19 before discharge and on the first day of admission.

|  | normal value | Admission day ( n=50 ) | Before discharge ( n=50 ) | P value |
| --- | --- | --- | --- | --- |
| WBC | 3.5-9.5 | 5.3 ( 4.0-7.7 ) | 5.7 ( 4.5-6.9 ) | 0.791 |
| N | 1.8-6.3 | 3.6 ( 2.5-5.8 ) | 3.6 ( 2.7-4.4 ) | 0.504 |
| L | 1.1-3.2 | 1.1 ( 0.6-1.6 ) | 1.5 ( 1.2-1.8 ) | 0.008 |
| HB | 120-160 | 136 ( 123-152 ) | 131 ( 118-140 ) | 0.042 |
| PLT | 125-350 | 191 ( 134-236 ) | 262 ( 194-332 ) | <0.001 |
| CK | 50-310 | 54 ( 38-81 ) | 36 ( 27-55 ) | <0.001 |
| LDH | 120-250 | 230 ( 187-274 ) | 187 ( 150-209 ) | <0.001 |
| AST | 15-40 | 21 ( 16-30 ) | 18 ( 15-26 ) | 0.421 |
| ALT | 9-50 | 18 ( 14-30 ) | 26 ( 17-42 ) | 0.033 |
| ALB | 40-55 | 40 ( 36-43 ) | 40 ( 36-41 ) | 0.067 |
| Scr | _* | 67 ( 60-79 ) | 70 ( 60-81 ) | 0.304 |
| BUN | 3.1-8 | 4.1 ( 3.3-5.6 ) | 3.8 ( 3.1-4.5 ) | 0.110 |
| D-D | 0-0.55 | 0.28 ( 0.19-0.50 ) | 0.40 ( 0.23-0.80 ) | 0.363 |
| CRP | 0-5 | 10.2 ( 2.7-43.8 ) | 2.6 ( 1.0-4.6 ) | <0.001 |
| PCT | <0.046 | 0.043 ( 0.032-0.068 ) | 0.037 ( 0.025-0.057 ) | 0.176 |
| IL-6 | 0-5.4 | 0 ( 0-8.4 ) | _<sup>△</sup> | 0.112 |

Comment : Because of the death of a critically ill patient, the impact was excluded, and only 50 cases were statistical. \*The standard values of serum creatinine in different age groups are different. <sup>△</sup>18 patients with interleukin-6 data lost at discharge because no tests were performed. All data are presented as medians (interquartile ranges, IQR).Laboratory indicator unit : HB g/l, CK U/L,LDH IU/L,AST U/L,ALT U/L,ALB g/l, Scr umol/l, BUN mmol/l, WBC 10<sup>9</sup>/L,N 10<sup>9</sup>/L,L 10<sup>9</sup>/L,PLT 10<sup>9</sup>/L,CRP mg/l, PCT ng/ml,IL-6 pg/ml, D-D mg/l. P<0.05 was considered statistically significant.

### Imaging examination

51 cases of chest CT were abnormal at the time of admission. During the course of the disease, chest CT mostly showed multiple patchy, flocculent fuzzy shadows, ground glass shadows or high-density shadows, some of which were consolidation; There were 41 cases of typical ground glass shadow (80.4%), 17 cases of local consolidation of pneumonia (33.3%), 20 cases of pleural thickening (39.2%), 7 cases of small amount of pleural effusion (13.7%), 8 cases of streak shadow of lung (15.7%), 4 cases of multiple solid nodular shadow (7.8%), 2 cases of air bronchogram (3.9%).2 cases (3.9%) had Single lobe lesions,49 cases (96.1%) had Multiple lobe lesions (Table 4). In addition to one dead patient, 50 patients were reexamined with chest CT within 2 days before discharge. The lesions increased in 1 case (2%), no significant change in 4 cases (8%), and decreased in 45 cases (90%).

**Table 4.** Radiographic(Abnormalities on chest CT)findings of 51 patients with COVID-19.

|  | N0.(%) |  |  | P value |
| --- | --- | --- | --- | --- |
|  | Total<br>(n=51) | Non-severe<br>(n=44) | Severe<br>(n=7) |  |
| Single lobe lesions | 2 ( 3.9) | 2 (4.5) | 0 ( 0 ) | 1.0 |
| Multiple lobe lesions | 49 ( 96.1) | 42(95.5) | 7(100) | 1.0 |
| Typical ground glass shadow | 41 ( 80.4) | 35 ( 79.5 ) | 6(85.7) | 0.703 |
| Local consolidation of pneumonia | 17( 33.3) | 12 ( 27.3 ) | 5(71.4) | 0.336 |
| pleural thickening | 20 ( 39.2) | 14 ( 31.8 ) | 6(85.7) | 0.007 |
| Small amount of pleural effusion | 7 ( 13.7) | 1 ( 2.3 ) | 6(85.7) | <0.001 |
| Lung streak shadow | 8( 15.7) | 8(18.2) | 0 ( 0 ) | 0.219 |
| Multiple solid nodules | 4 (7.8) | 4(9.1) | 0 ( 0 ) | 1.0 |
| Bronchial shadow with air | 2 (3.9) | 1 ( 2.3 ) | 1 ( 14.3 ) | 0.258 |

Comment : All data are presented as N0.(%). P values denoted the comparison between non-severe cases and severe cases. P<0.05 was considered statistically significant.

### Treatment

According to their clinical manifestations and traditional Chinese medicine(TCM) symptoms, most patients were given antiviral treatment with TCM Decoction (28 [54.9%]);All of them received aerosol inhalation of recombinant human interferon a-1b for injection and oral antiviral therapy with Lopinavir and Ritonavir tablets (51 [100%]), some patients were treated with oseltamivir (7 [13.7%]), and 2 patients (3.9%) were given antiviral therapy with arbidol after discontinuation of Lopinavir and Ritonavir tablets;Most of the patients were given Bacillus licheniformis capsules regulated intestinal flora treatment (44 [86.3%]); 48 cases (94.1%) were treated with thymopentin for injection;Most of the patients with fever were treated with reduning injection, clearing away heat, dispersing wind and detoxifying (30 [58.8%]);11 patients (21.6%) were treated with antibiotics; 10 patients (19.6%) received short-term (3-5 days) glucocorticoid treatment(Table 5).

**Table 5.**
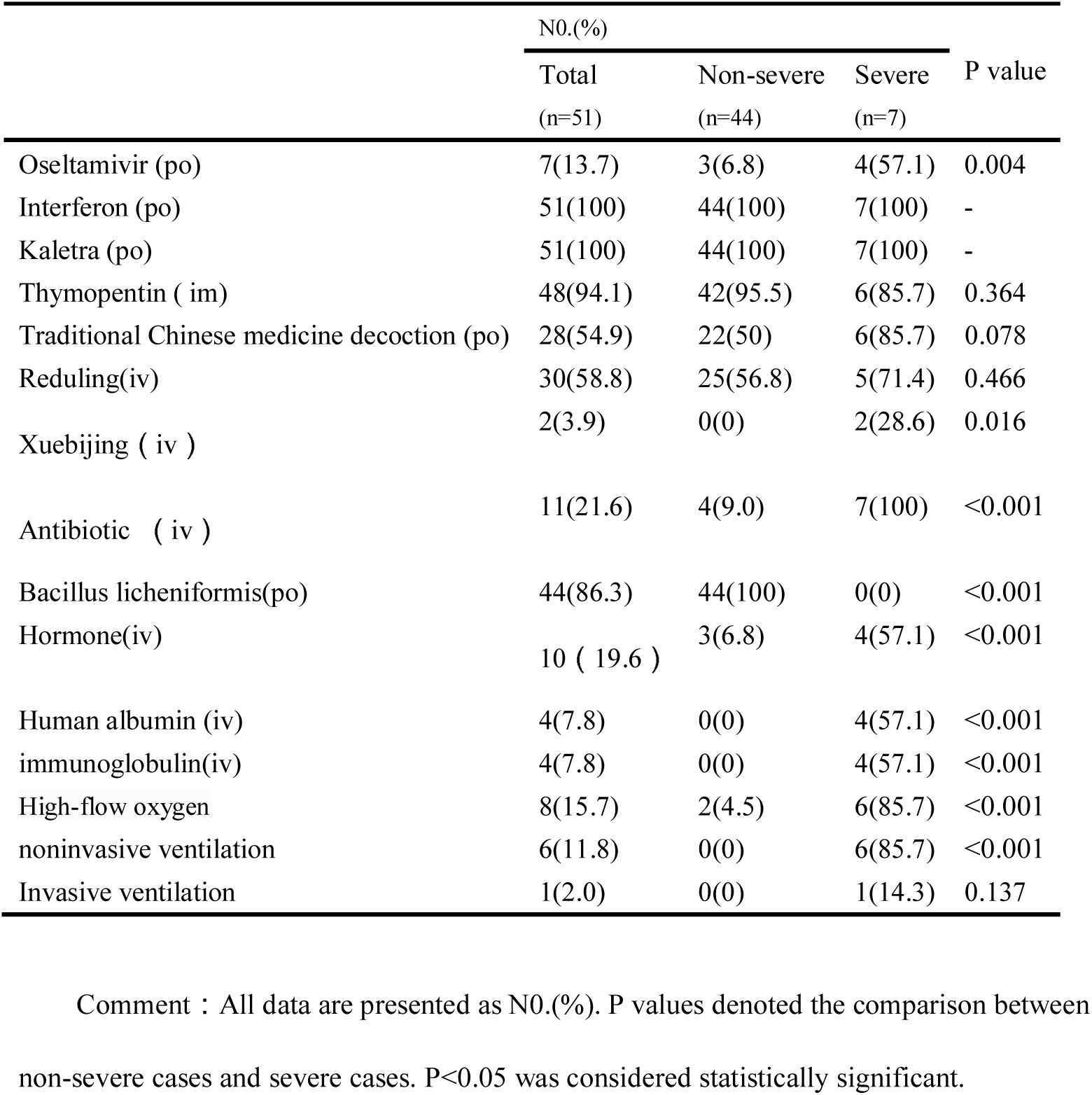
Treatment of 51 patients with COVID-19.

In 7 severe patients, 6 patients(85.7%) had obvious appetite decline, and they were given “short peptide enteral nutrition solution, protein component diet, dietary fiber component diet” nutritional support treatment; in 7 severe patients, 4 patients (57.1%) were given human albumin infusion treatment, 4 patients (57.1%) were given immunoglobulin treatment(Table 5).

### Comparison of clinical characteristics between severe patients and non-severe patients

Compared with non-severe patients (n = 44), severe patients (n = 7) were older (median age, 52 years vs 44 years), had a higher proportion of diabetes mellitus (4 [57.1%] vs 0 [0.0%]), had a lower lymphocyte levels (0.37[0.3-0.6] vs 1.3[0.9-1.7]),had a higher CRP levels(135.7[83.9-180.9] vs 7.5[2.4-23.8]),most of them had a small amount of pleural effusion (6 [85.7%] vs 0 [2.3%]),most of them needed antibiotic treatment (7 [100%] vs 4 [9.1%], most of them needed nutritional diet (6 [85.7%) vs 0 [0.0%], and were more likely to have dyspnea (6 [85.7%] vs 5 [11.4%]),most of them needed noninvasive mechanical ventilation (6 [85.7%] vs 0 [0.0%])(Table 1,2,4,5). ARDS was found in 3 severe patients, and 1 patient died with shock complications.

### Clinical outcomes

Except one patient died, the remaining 50 patients were discharged according to the discharge standard, the common clinical symptoms disappeared basically, the lymphocyte increased significantly (P =0.008), CRP decreased significantly (P <0.001). The median length of stay was 12 days (IQR, 9-13).

## Discussion

This study is a series of patients who have been discharged from non-Wuhan area. As of February 11, 2020, a total of 51 patients were discharged from the single center of the Chongqing University Three Gorges Hospital, of which 50 were cured and discharged, and 1 critically Patients was died. Among the 51 patients in this study, the median number of days from onset to Confirmed diagnosis of COVID-19 was 6 days (IQR, 3-8 days); among the 50 patients who were discharged alive, the Average hospitalization day was 12 days (IQR, 9-13 days).

Most patients had visited Wuhan or other Hubei areas or had close contact with COVID-19 patients within 14 days before the onset of the disease, but 4 cases had no clear epidemiological history of COVID-19 patients, which indicates that there may be active SARS-CoV-2 transmission. The rapid spread of close contact between people is an important feature of COVID-19 [12], which requires close monitoring to prevent further large-scale spread of SARS-CoV-2 worldwide. The 51 infected people were mainly young adults, mostly imported cases, which may be related to large-scale population migration during the Chinese New Year.

The more common clinical symptoms in this group of cases are: fever 43 (84.3%), cough 38(74.5%)and fatigue 22 (43.1%). However, a small proportion of patients initially did not feel obvious fever, atypical Symptoms, such as diarrhea and nausea and retching, are similar to Professor Zhong Nanshan’s study[13].One patient in this group Without any discomfort and clinical symptoms, was Be hospitalized for a history of epidemiology.

On the first day of admission, the decrease of lymphocyte count was more common (26 [51.0%]); the majority of leukocytes and neutrophils were in the normal range; most patients (32 [62.7%]) had an increase of CRP; some patients had an increase of LDH (20 [39.2%]), D-dimer (10 [19.6%]), IL-6 (16 [31.4%]) and PCT (25 [49%]); and urine routine urine protein was positive in 8 cases. Compared with the non-severe patients, the lymphocyte count decreased more significantly in the severe patients, the serum albumin level was lower, and the LDH, CRP, D-dimer and PCT levels were significantly higher. Compared with the results of various laboratory indexes of patients on the first day of admission (N=50) and before discharge (N=50), we found that the lymphocyte and PLT of patients before discharge increased significantly, while CK, LDH and CRP decreased significantly.

Therefore, we speculate that a decrease in lymphocyte count and an increase in CRP are an important feature of COVID-19. Compared with non-severe patients, severe patients have a lower lymphocyte count and an increase in CRP. The lymphocyte count was significantly increased and CRP was significantly decreased before discharge of the 50 surviving patients,which suggested that the decrease of absolute lymphocyte count and the increase of CRP might be related to the severity of the disease. Therefore, dynamic observation of lymphocyte count and CRP level may be of great significance to the change of the disease and prognosis of the patients, which needs further study and confirmation.

51 cases of chest CT were abnormal at the time of admission. During the course of the disease, chest CT mostly showed multiple patchy, flocculent fuzzy shadows, ground glass shadows or high-density shadows, some of which were consolidation; most of them had typical ground glass Shadows (41 cases [80.4%]);In severe patients (n = 7), most of them had pleural thickening (6 [85.7%]) and a small amount of pleural effusion (6 [85.7%]), while non-severe patients,the proportion is smaller (14 [31.8%], 1 [2.3%]). This suggests that in the course of COVID-19 clinical treatment, if chest CT indicates a small amount of pleural effusion with pleural thickening, the patient may be in a serious condition, which needs to be paid attention to and actively treated.

The common clinical symptoms of 50 living patients basically disappeared at the time of discharge, but the chest CT lesions still existed. The lesions increased in 1(2%) case, no significant change in 4 (8%)cases, and decreased in 45 (90%)cases. This suggests that the imaging manifestations of chest may not be parallel to the clinical symptoms. Lung lesions still exist after the negative detection of SARS-CoV-2 nucleic acid test. Further follow-up is needed.

Previous studies on SARS and MERS [19] showed that,antibacterials and antiviral drugs had no definite effect on SARS and MERS. In a recent study[12],all of the patients with COVID-19 received antibacterial agents, 90% received antiviral therapy, and 45% received methylprednisolone. However, no effective outcomes were observed.There is no clear evidence that patients with SARS-CoV-2 infection will benefit from hormone therapy and are more likely to face the risks of hormone therapy. Glucocorticoid is not recommended for the treatment of COVID-19[20].

Therefore, traditional Chinese medicine (TCM)treatment may be a useful choice. Although Western medicine is currently used in many Eastern countries as a mainstream medicine, Chinese medicine also plays an important role against various diseases. As we know, to reach a conclusion regarding the effectiveness of Chinese medicine in any specific disease, controlled clinical trials are needed. Moreover, for a devastating epidemic outbreak like SARS, only retrospective study reports can be found [21]. Chinese medicine may also have a role in the prevention of acute and critical diseases like the SARS virus. Chinese medicine was used as adjuvant therapy in the SARS patients. It seems that the combined treatment(Chinese herbs combined with western medicine)was helpful for SARS patients. Chinese herbs combined with Western medicines made no difference in decreasing mortality versus Western medicines alone. It is possible that Chinese herbs combined with Western medicines may improve symptoms, quality of life and absorption of pulmonary infiltration, and decrease the corticosteroid dosage for SARS patients [22].

Among the 51 patients in this study, Except for routine administration of recombinant human interferon a-1b for injection nebulized inhalation, oral antiviral therapy with lopinavir or ritonavir tablets (51 [100%]), According to their clinical manifestations and TCM syndromes, most patients were given antiviral therapy with Chinese medicine decoction (28 [54.9%]), and achieved good clinical effects. But just like above-mentioned, to reach a conclusion regarding the effectiveness of Chinese medicine in COVID-19,controlled clinical trials are needed.

Compared with non-severe patients (n = 44), severe patients (n = 7) were older (median age, 52 years vs 44 years), with a higher proportion of diabetes mellitus (4 [57.1%] vs 0 [0.0%]);This suggests that age and comorbid diabetes may be an important factor affecting the patient’s condition and prognosis, which was also confirmed by Wei J’s study [12]. In this study, only one critically ill patient (dead patient) needs invasive mechanical ventilation, which is lower than previous studies in Wuhan [1, 12]. Due to the shortage of early medical resources in Wuhan [13, 14], this may be related to the serious condition of inpatients in Wuhan.

This study has several limitations. First, the sample size is small, especially in severe patients. Second, the majority of discharged patients are non-severe patients. Severe and critically ill patients often have more complications and may require longer hospital stays. Most are still in hospital. Third, there are four COVID-19 treatment centers in Chongqing, and there is still no relevant comparative analysis.

## Conclusions

In the case analysis of 51 discharged patients of COVID-19 in the single center of Northeast Chongqing, most of the infected patients were young and middle-aged, and most of them were imported cases. Most cases have typical symptoms such as fever and cough, and a few cases have atypical symptoms such as diarrhea. The changes of CRP and other inflammatory indexes may be related to the severity of the disease. Traditional Chinese medicine provides an alternative for the treatment of COVID-19.

## Data Availability

The datasets generated and analyzed during the current study are available from the corresponding author on reasonable request.

## Acknowledgement

We sincerely thank Lian Guo, director of the Teaching Department of the Chongqing Three Gorges Central Hospital, and Chunhui Lang, director of the Foreign Affairs Department of Scientific Research, for their great support to this subject.

## Author’s contribution

L.L. and J.G. had the idea for and designed the study and had full access to all data in the study and take responsibility for the integrity of the data and the accuracy of the data analysis.

L.L, J.G., W.H. and C.L. contributed to writing of the report.

L.G., C.L. and Y.T. contributed to critical revision of the report.

J.G., L.L. and Q.P. contributed to the statistical analysis.

All authors contributed to data acquisition, data analysis, or data interpretation, and reviewed and approved the final version

## References

1. Huang C, Wang Y, Li X, et al. Clinical features of patients infected with 2019 novel coronavirus in Wuhan, China [published online ahead of print, 2020 Jan 24]. Lancet. 2020; DOI:10.1016/S0140-6736(20)30183-5.

2. WHO Director-General’s remarks at the media briefing on 2019-nCoV on 11 February 2020. Retrieved February 11, 2020, from https://www.who.int/dg/speeches/detail/who-director-general-s-remarks-at-the-media-briefing-on-2019-ncov-on-11-february-2020.

3. Gorbalenya. (2020). Severe acute respiratory syndrome-related coronavirus – The species and its viruses, a statement of the Coronavirus Study Group. Retrieved bioRxiv, https://doi.org/10.1101/2020.02.07.937862.

4. International Committee on Taxonomy of Viruses. Retrieved February 11, 2020, from https://talk.ictvonline.org/.

5. WHO main website. https://www.who.int (accessed February 14th, 2020)

6. Phan LT, Nguyen TV, Luong QC, et al. Importation and human-to-human transmission of a novel coronavirus in Vietnam. N Engl J Med. 2020; doi: 10.1056/NEJMc2001272

7. Rothe C, Schunk M, Sothmann P, et al. Transmission of 2019-nCoV infection from an asymptomatic contact in Germany. N Engl J Med. 2020; DOI:10.1056/NEJMc2001468

8. Holshue ML, DeBolt C, Lindquist S, et al. First case of 2019 novel coronavirus in the United States. N Engl J Med. 2020; doi: 10.1056/NEJMoa2001191

9. William Kyle Silverstein, Lynfa Stroud, Graham Edward Cleghorn, et.al. First imported case of 2019 novel coronavirus in Canada, presenting as mild pneumonia. The Lancet; February 13, 2020. DOI:https://doi.org/10.1016/S0140-6736(20)30370-6

10. National Health Commission of the People’s Republic of China. http://www.nhc.gov.cn (Assessed on February 15th, 2020)

11. Chen N, Zhou M, Dong X, et al. Epidemiological and clinical characteristics of 99 cases of 2019 novel coronavirus pneumonia in Wuhan, China: a descriptive study. Lancet. 2020. DOI:10.1016/S0140-6736(20)30211-7.

12. Wang D, Bo Hu B,Hu C, et al. Clinical Characteristics of 138 Hospitalized Patients With 2019 Novel Coronavirus–Infected Pneumonia in Wuhan, China. JAMA. February 2020.DOI: 10.1001/jama.2020.1585

13. Wei J, Ni Z, Hu-Y, et al. Clinical characteristics of 2019 novel coronavirus infection in China. Medrxiv, Posted February 09, 2020. doi : https://doi.org/10.1101/2020.02.06.20020974

14. Yang Y, Lu Q, Liu M, et al. Epidemiological and clinical features of the 2019 novel coronavirus outbreak in China. Medrxiv, Posted February 11, 2020.doi: https://doi.org/10.1101/2020.02.10.20021675

15. Chongqing Health Committee. http://wsjkw.cq.gov.cn (Assessed on February 15th, 2020)

16. World Health Organization. Clinical management of severe acute respiratory infection when novel coronavirus (nCoV) infection is suspected: interim guidance. Published January 28, 2020. Accessed January 31, 2020. https://www.who.int/publications-detail/clinical-management-of-severe-acute-respiratory-infection-when-novel-coronavirus-(ncov)-infection-is-suspected

17. The national health and Health Commission of the people’s Republic of China, State Administration of traditional Chinese medicine of the people’s Republic of China. Diagnosis and treatment of novel coronavirus pneumonia (trial version fifth). 2020.

18. Ranieri VM, Rubenfeld GD, Thompson BT, et al. ARDS Definition Task Force. Acute respiratory distress syndrome: the Berlin definition. JAMA.2533-2526:(23)307;2012doi/10.1001:jama.2012.5669.

19. De Wit E, Van Doremalen N, Falzarano D, et al. SARS and MERS: recent insights into emerging coronaviruses. Nat Rev Microbiol. 534-523:(8)14; 2016. doi/10.1038:nrmicro.2016.81PubMedGoogle ScholarCrossref.

20. Clark D Russell, Jonathan E Millar, J Kenneth Baillie, et al. Clinical evidence does not support corticosteroid treatment for 2019-nCoV lung injury. Published:February 11, 2020.https://doi.org/10.1016/S0140-6736(20)30317-2

21. Luo Y, Wang C Z, Hesse-Fong J, et al. Application of Chinese Medicine in Acute and Critical Medical Conditions. The American Journal of Chinese Medicine, 2019:1–13.

22. Liu X, Zhang M, He L, et al. Chinese herbs combined with Western medicine for severe acute respiratory syndrome (SARS). Cochrane Database of Systematic Reviews. 2012, 10 (10), CD004882. DOI:10.1002/14651858.CD004882.pub3

